# Occupational inequalities in the prevalence of COVID-19: A longitudinal observational study of England, August 2020 to January 2021

**DOI:** 10.1101/2021.06.01.21258140

**Authors:** Mark A. Green, Malcolm G. Semple

## Abstract

**Background:** The COVID-19 pandemic has reinforced, amplified and created new health inequalities. There is less evidence on how COVID-19 prevalence varies by measures of work and occupation which represent a key social determinant of health. The aim of the study is to evaluate how occupational inequalities in the prevalence of COVID-19 varies across England and their possible explanatory factors.

**Methods:** We used data for 363,651 individuals (2,178,835 observations) aged 18 years and over between 1st May 2020 and 31st January 2021 from the ONS Covid Infection Survey, a representative longitudinal survey of individuals in England. We focus on two measures of work; employment status for all adults, and work sector of individuals currently working. Multi-level binomial regression models were used to estimate the likelihood of testing positive of COVID-19, adjusting for known explanatory covariates.

**Results:** 0.9% of participants tested positive for COVID-19 over the study period. COVID-19 prevalence was higher among adults who were students or furloughed (i.e., temporarily not working). Among adults currently working, COVID-19 prevalence was highest in adults employed in the hospitality sector, with higher prevalence for individuals employed in transport, social care, retail, health care and educational sectors. Inequalities by work were not consistent over time.

**Conclusions:** We find an unequal distribution of infections relating to COVID-19 by work and employment status. Our findings demonstrate the need for greater workplace interventions to protect employees, but also that a large proportion of SARS-CoV-2 transmission occurs outside of work. In particular, populations who experienced social and economic harms through being furloughed were also more likely to experience a double burden of increased likelihood of COVID-19.

## Introduction

The social, health and economic impacts resulting from the spread of Severe Acute Respiratory Syndrome Coronavirus-2 (SARS-CoV-2), and restrictions aimed at managing its spread, have been unprecedented in scale and scope. In England, as in most countries, the impacts of Coronavirus disease 2019 (COVID-19) resulting from SARS-CoV-2 have been unevenly felt across populations. Infection, hospitalisation and mortality outcomes have been higher in older populations, males, Black and Asian ethnic groups, and deprived communities (Docherty *et al*., 2020; Platt and Warwick, 2020; Public Health England, 2020; Harrison *et al*., 2021; HM Government, 2021b; Whitehead, Taylor-Robinson and Barr, 2021). Understanding and tackling the social inequalities arising from and amplified by COVID-19 remains a core government priority.

As our personal and social lives have had to adapt to the COVID-19 pandemic, so too has our economic, work and employment circumstances to minimise transmission of SARS-CoV-2. Many employment roles were ‘furloughed’ (i.e., temporary unemployment), with salaries being covered by the government if an employer did not make them unemployed. Some occupation roles adapted so that individuals could work from home, whereas others were able to introduce protective social distancing measures into work. However, not all occupations were able adapt to either of these strategies. Emerging evidence has demonstrated that ‘essential’ occupations who work directly with patients (e.g., health or social care workers), groups unable to work from home (e.g., transport or manufacturing occupations), or occupations with ‘front facing’ roles where individuals are routinely exposed to others (e.g., supermarket workers or teachers) were at greater risk of COVID-19 and associated severe outcomes including mortality (de Gier *et al*., 2020; ONS, 2020, 2021a; Chen *et al*., 2021; Gholami *et al*., 2021; HM Government, 2021a; Mutambudzi *et al*., 2021). Many of these occupational risk factors also intersect with age, sex, ethnicity and deprivation, partly explaining or amplifying health inequalities (Paremoer *et al*., 2021; Whitehead, Taylor-Robinson and Barr, 2021). As such, the ability of work to adapt or change within government restrictions was experienced unevenly across the population and may partly explain the pathways through which social inequalities in COVID-19 have materialised. Identifying work sectors at highest risk is key for designing social distancing and preventative interventions to help re-open society during the roll-out of vaccines (Brooks-Pollock *et al*., 2021).

The aim of this study is to evaluate how occupational inequalities in the prevalence of COVID-19 vary across England and their possible explanatory factors. We address several limitations in the literature investigating this issue. First, there is a paucity of evidence on the extent of risks of COVID-19 by occupational groups for all ages. We need to better understand how risk varies across these groups to better develop interventions to minimise transmission risk for managing COVID-19 and future pandemics. Second, evidence derived from ‘testing’ data are biased due to self-selection (i.e., focusing on individuals with symptoms). We tackle this through using a novel survey where participants were all tested irrespective of symptoms. Third, our large survey helps accommodate issues relating to the rarity of events, allowing for detailed investigations into occupational inequalities in COVID-19.

## Methodology

### Data

The ONS Covid Infection Survey (CIS) was used as the primary data source. The CIS is a representative random sample survey of the population in England used to monitor trends in COVID-Individuals are invited take a COVID-19 test irrespective of whether they have symptoms or not, allowing an estimate of overall COVID-19 prevalence. Individuals also complete a survey about a range of demographic, social and health questions that contextualise their circumstances. There were 2,772,698 observations between 1st May 2020 and 31st January 2021 available for analysis.

Data from August onwards (n = 2,518,142) were selected to assess trends during the second wave of infections in England. While the CIS started in May, with low infection levels and limited data collection between May and August, these data were removed. Although August also has low levels of infections, it was included to capture the baseline data and seek signals preceding the star the second wave occurring in September. Only observations for adults aged 18 years and over were selected for the analysis (n = 2,178,835).

While the CIS is a repeated cross-sectional survey, individuals were encouraged to take part repeatedly over time. Participants were asked to enrol for follow-up waves, initially weekly over the first month then monthly up to 13 times (ONS, 2021b). Our study leverages this longitudinal design. The analytical sample contained 363,651 individuals. 61% of individuals had at least one record per month (mean number of records over the study period was 6, with a standard deviation of 2.2). 5% of individuals had only one occurrence in the data. We utilise each observation as the primary unit of analysis, nested within individuals.

The outcome variable for our analysis was whether an individual had tested positive for COVID-19 or not (binary). Tests were nose and throat swabs self-administered by participants and posted for analysis at a hub laboratory. Swabs were tested for SARS-CoV-2 using reverse transcription polymerase chain reaction (PCR) tests (ONS, 2021b). Tests recorded as ‘void’ or ‘insufficient’ were excluded from the analysis.

We selected two measures of occupation and work as our primary measures of interest. First, *employment status* was chosen to represent an individual’s primary employment circumstances as an aggregate measure of work-related risk. Categories were employed, self-employed, furloughed (i.e., individuals temporarily not working), student, or not working (e.g., retired, economically inactive, unemployed). Second, focusing on just individuals who are currently working either employed or self-employed, we also consider *work sector*. 15 categories of occupational sectors (e.g. teaching and education, health care, retail sector) were used to assess differences in COVID-19 risk by type of work. Low sample sizes for specific occupations meant they were less suitable for the analysis. Descriptions of each work sector can be found in Appendix Table A.

Additional explanatory variables were selected based on key factors that may help to explain occupational differences in COVID-19 to adjust for them in our analyses:

- *Age* – included to assess differences in risk by age, due to evidence that younger population groups were more likely to have tested positive for COVID-19 and older age groups more likely to have experienced severe harms relating to COVID-19 (Docherty *et al*., 2020; Public Health England, 2020). Age is used both in its raw value, as well as squared to account for possible non-linear effects.
- *Sex* – included to assess the differences in factors affecting males and females differently (Docherty *et al*., 2020; Public Health England, 2020).
- *Ethnicity* - selected due to inequalities in social and health impacts of COVID-19 disproportionally affecting non White British populations (Platt and Warwick, 2020; Public Health England, 2020; Harrison *et al*., 2021; HM Government, 2021b). Ethnic groups were kept as specific groups where possible, although some groups were combined together to ensure sufficient sample sizes and avoid data disclosure issues.
- *Number of people within a household* – chosen as a greater number of people may increase opportunities for the spread of COVID-19 (Madewell *et al*., 2020).
- *Whether an individual had travelled abroad recently or not* – include due to the possible higher risk from individuals travelling to countries with higher COVID-19 prevalence or greater social mixing (Russell *et al*., 2021).
- *Work location* – for analyses using work sector only, we account for whether individuals were working at home, outside the home or a mixture of both. Individuals who are working outside of the home may have higher risk as they may be mixing with other individuals or have greater exposure to SARS-CoV-2 (HM Government, 2021a).
- *Geographical location* – region of England an individual was resident in to account for the spatial heterogeneity in COVID-19 (Public Health England, 2020). Regions were created by the ONS and match Local Authority districts, with districts combined to make sure no region has a population of less than 500,000 to preserve data security. There were 116 regions.
- *Month* – we also adjust for month of the year to account for the differential risk of COVID-19 that varied over time, although do not report these results.

### Statistical analyses

Descriptive summary statistics and visualisations are used to describe aggregate patterns in the data. We first describe demographic patterns in our outcome variable to help contextualise our statistical analyses. Multi-level binomial regression models were used to analyse the risk of COVID-19. Models control for a series of fixed effects representing individual-level covariates that may explain differences in COVID-19 risk. Two random effects are included: (i) participant ID (varying intercept) to account for repeat observations within the survey over time, and (ii) geographical area (varying intercept) to account for geographical inequalities in COVID-19 prevalence and therefore risk. Numeric values were z-score standardised to minimise issues with their different scales (age and household size).

We map the conditional mean estimated from the model to aid interpretation of how COVID-19 likelihood varied spatially. Two types of maps are used. First, conventional geographical zones are used. Second, ‘hexmaps’ are also used to aid interpretation. While the hexmap is more abstract, all areas are given equal size (arranged as close to match the geographical pattern as possible) therefore minimising any distorted patterns created through rural areas being larger in size than urban areas despite smaller populations.

## Results

### Demographic inequalities in COVID-19 prevalence

Summary sample characteristics can be viewed in the Appendix (see Tables B and C). 0.9% of respondents tested positive for COVID-19 during the study period. Figure 1 presents trends in the estimated prevalence of COVID-19 during the study period. Following low levels of COVID-19 in August, prevalence of COVID-19 began to rise in September onwards peaking in the first week of November. COVID-19 prevalence declines thereafter, following a national lockdown on 5th November 2020, before rising again after the end of the lockdown (2nd December 2020) and with the emergence of the B1.1.7 variant. Trends then decline following the national lockdown announced on the 6th January 2021. There were no noticeable differences in trends between males and females.

**Figure 1:**
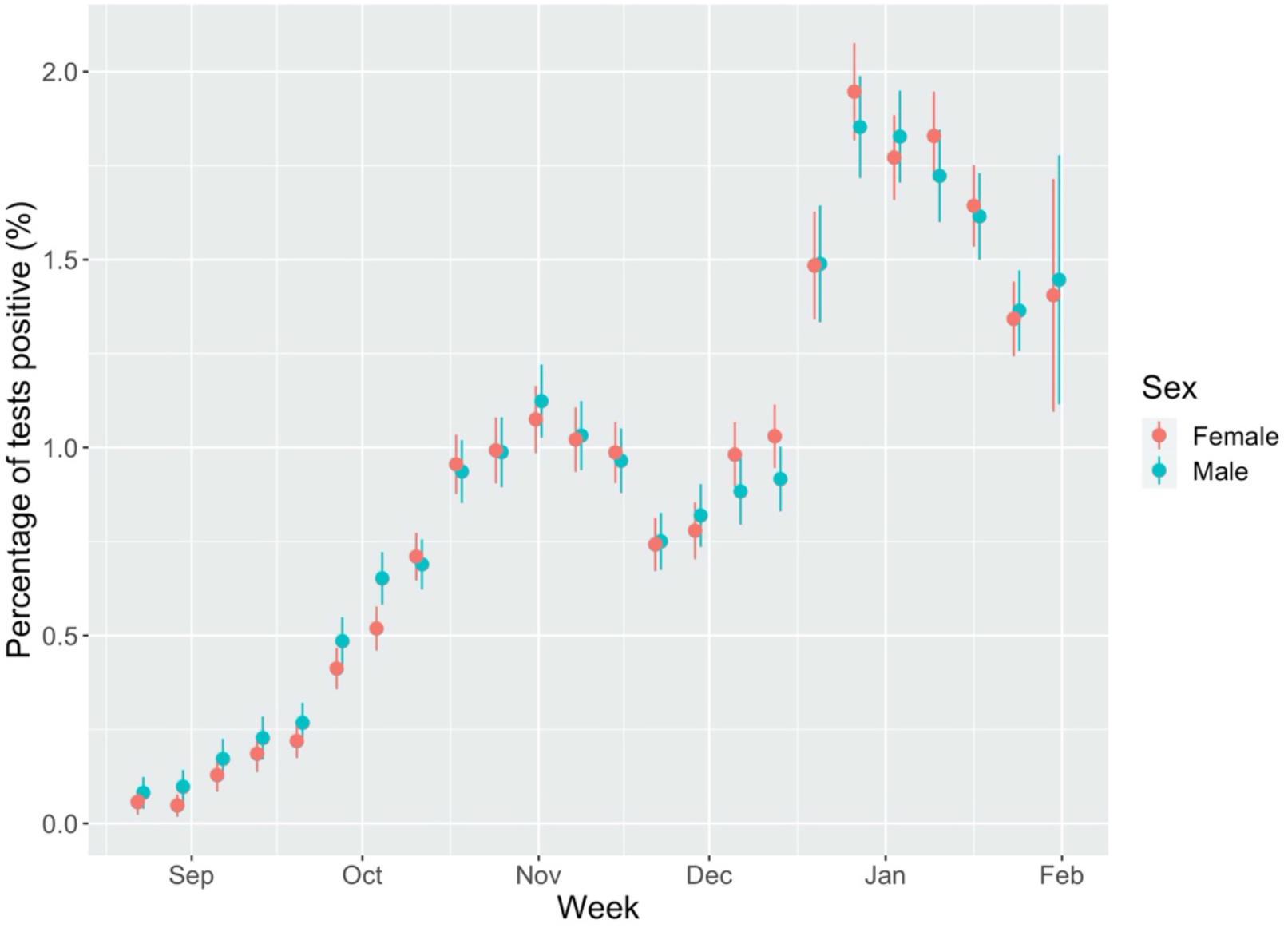
**Percentage of tests that were positive for COVID-19 by week of year and sex. (Note: figures in first two weeks of August were redacted due to disclosive numbers (i.e., <10 positive tests. Point is estimated percentage, with error bars the 95% confidence intervals)**.

Figure 2 examines how COVID-19 prevalence varies by single year of age. Highest prevalence of COVID-19 was among ages 18-22 with prevalence at least twice as high as the national average. Prevalence was higher among these ages for females compared to males, although there were overlapping confidence intervals limiting the conclusions we can draw here. Prevalence of COVID-19 declines therefore with age thereafter.

**Figure 2:**
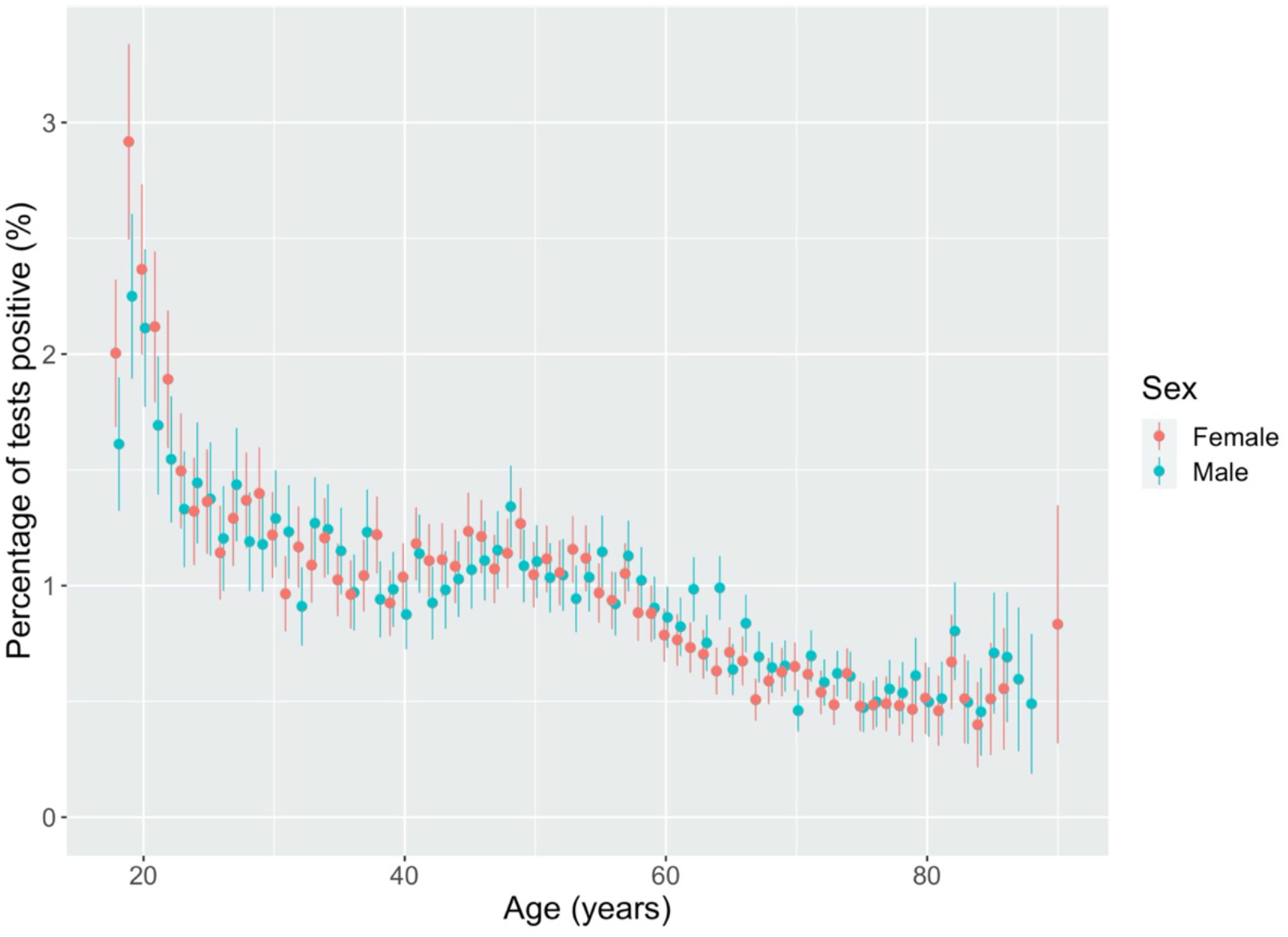
COVID-19 prevalence by age and sex.

Figure 3 presents COVID-19 prevalence by ethnic group. Highest prevalence of COVID-19 was found for Pakistani ethnicity (more than two times higher than the prevalence of the White British group), with higher prevalence also among Black African groups. Lower prevalence was observed for Chinese, White British, and Mixed White and Asian groups. There were no significant differences by sex.

**Figure 3:**
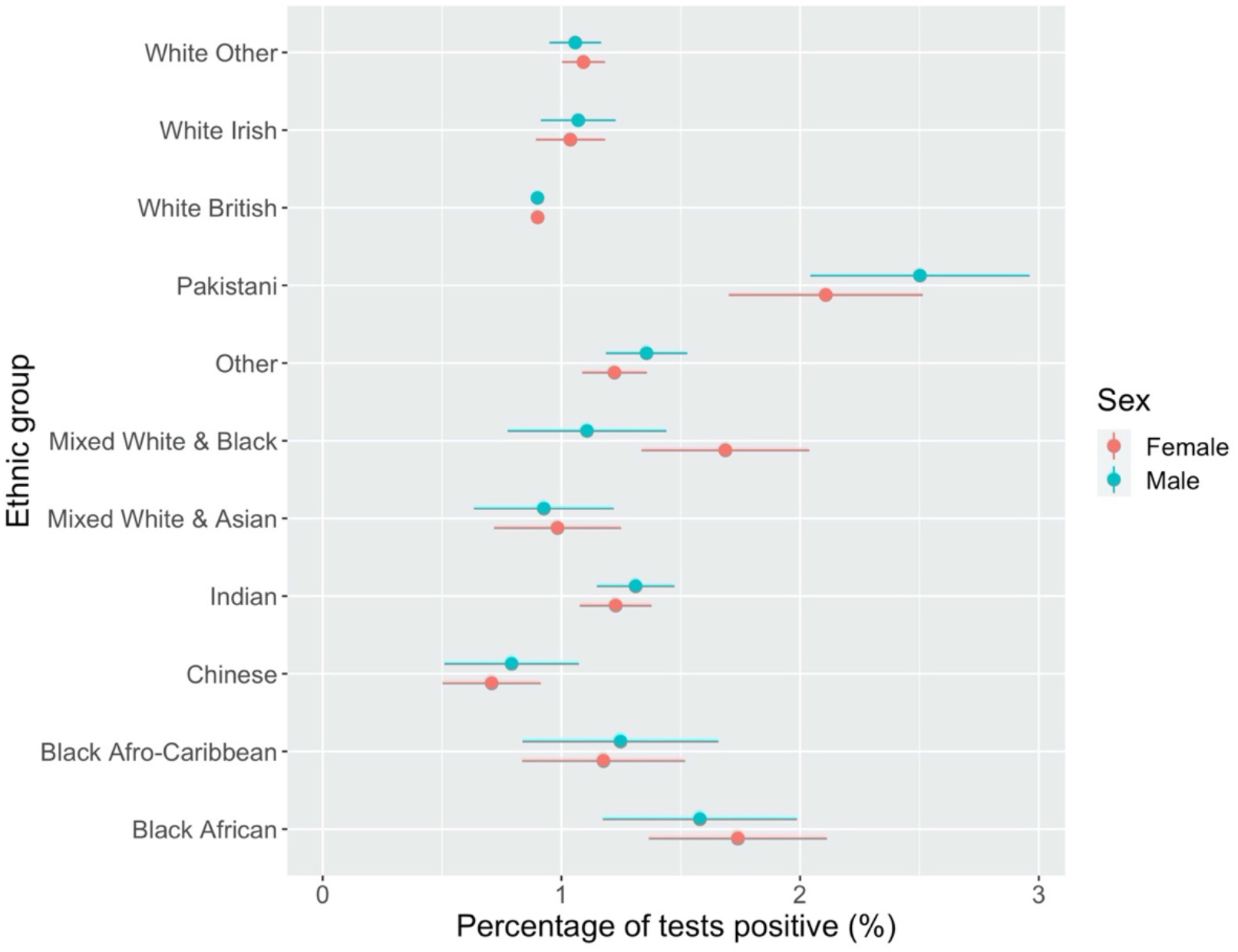
COVID-19 Prevalence by ethnic group and sex.

### Descriptive inequalities by employment type and work sector

We next consider the prevalence of COVID-19 by our two key measures of work and occupation. Figure 4 presents COVID-19 prevalence among all adults by employment status (Figure 4). The highest prevalence of COVID-19 was observed for individuals who were students or furloughed (i.e., individuals temporarily not working). Lower prevalence was estimated for individuals who were not working (i.e., retired, unemployed, long-term sick) and self-employed groups. There were no differences by sex. Stratifying analyses by age revealed higher prevalence of COVID-19 among younger populations for each employment type, especially younger furloughed males (see Appendix Figure A).

**Figure 4:**
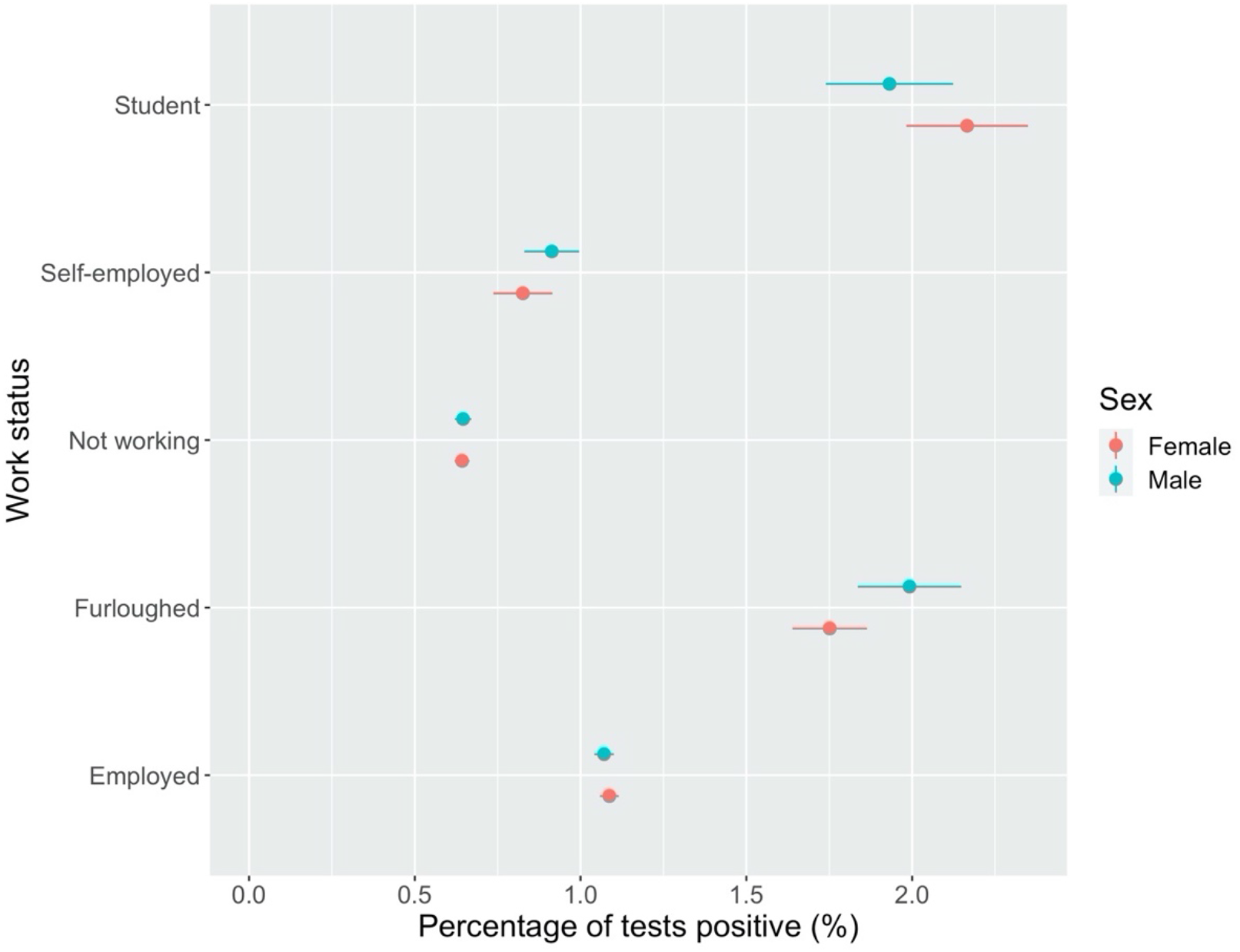
COVID-19 Prevalence by work status and sex.

We next consider inequalities in prevalence of COVID-19 by work sector (Figure 5). The highest prevalence of COVID-19 overall was found for individuals employed in the hospitality sector, with higher prevalence for individuals employed in transport, social care, retail, health care and education. Lowest prevalence was for individuals employed in ICT, with low prevalence in the armed forces and entertainment sectors. There were some differences in estimates between males and females, with higher prevalence among males for transport, retail, manufacturing, food production and hospitality. Higher prevalence for females was observed for social care and personal services. These may reflect sectors where males or females end up in different roles. However, limited conclusions can be drawn since confidence intervals overlapped for males and females for all work sectors other than retail and manufacturing, where prevalence was higher for males in both. Stratifying analyses by age showed higher risks across most work sectors for younger populations (Appendix Figure B).

**Figure 5:**
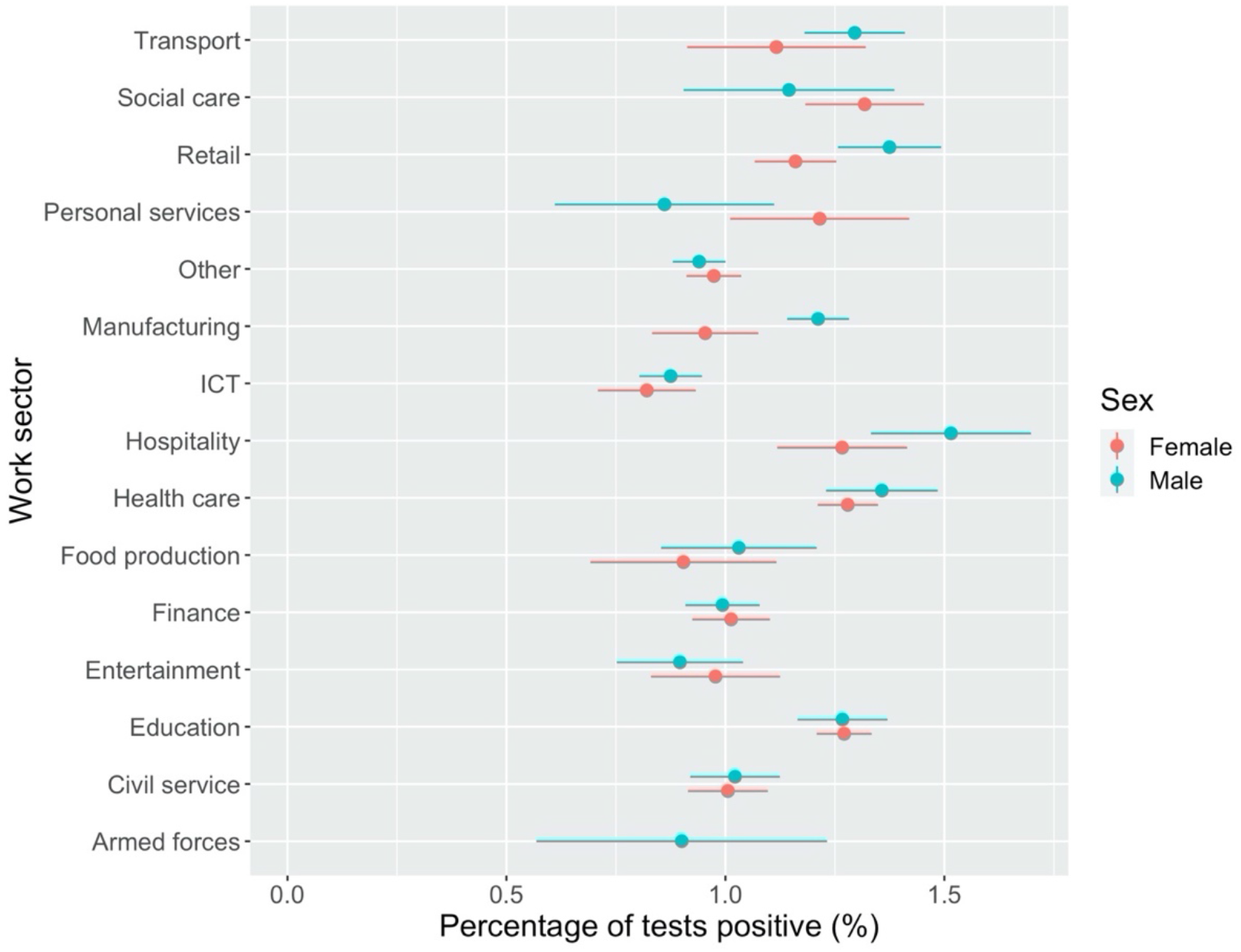
**COVID-19 prevalence by work sector and sex. Note: estimate for females employed in the armed forces excluded due to counts <10 to preserve ONS data disclosure standards**.

### Regression analyses of occupational risk of COVID-19

First, we examine for all adults the likelihood of having tested positive for COVID-19 by employment status (Table 1). In comparison to individuals who were employed, individuals who were furloughed were 81% more likely (Odds Ratio (OR) = 1.81, 95% Confidence Intervals (CIs) = 1.69-1.93) to have tested positive for COVID-19. Students were 35% (OR = 1.35, 95% CIs = 1.22-1.50) more like than employed individuals to test positive for COVID-19.

**Table 1:** **Model summary for analysing COVID-19 risk by socio-demographic features including occupational status. Note: Model also adjusted for time (month). Results placed in bold to emphasise associations where 95% confidence intervals (CIs) do not contain 1**.

| Variable | Odds Ratio | Lower CI | Upper CI |
| --- | --- | --- | --- |
| Male | Reference |  |  |
| Female | 0.994 | 0.958 | 1.031 |
| Age (z-score) | **0.858** | **0.763** | **0.965** |
| Age-squared (z-score) | 1.003 | 0.884 | 1.138 |
| White British | Reference |  |  |
| Any other ethnic group | 1.102 | 0.985 | 1.235 |
| Any other white background | 0.987 | 0.903 | 1.080 |
| Chinese | 0.624 | 0.468 | 0.833 |
| Indian | **1.130** | **1.004** | **1.273** |
| Pakistani | **1.687** | **1.389** | **2.048** |
| Black African | **1.359** | **1.086** | **1.700** |
| Black Afro-Caribbean | 1.123 | 0.848 | 1.486 |
| Mixed White & Asian | 0.874 | 0.675 | 1.131 |
| Mixed White & Black | 1.190 | 0.947 | 1.496 |
| White Irish | **1.204** | **1.025** | **1.415** |
| Employed | Reference |  |  |
| Self-employed | 0.927 | 0.851 | 1.011 |
| Furloughed | **1.808** | **1.692** | **1.933** |
| Not working | 0.834 | 0.790 | 0.880 |
| Student | **1.350** | **1.216** | **1.500** |
| Have travelled abroad recently | **1.132** | **1.072** | **1.196** |
| Household size (z-score) | **1.124** | **1.104** | **1.145** |
| Random effects | Variance | SD |  |
| ID (participant) | 3.149 | 1.774 |  |
| Geographical area | 0.130 | 0.360 |  |

There were also distinct demographic inequalities. Age was negatively associated with COVID-19 risk, so that older populations were less likely to have tested positively. No association for sex was detected. Ethnic inequalities were evident, with greater risk of COVID-19 found for Indian (13% more likely), Pakistani (69%), Black African (36%) and White Irish (20%) populations than compared to White British populations. We observed a greater likelihood of COVID-19 among individuals who had travelled abroad recently. Finally, there was a positive association to number of people in the household, suggesting greater prevalence of COVID-19 among larger households.

Table 2 presents our second model which considers only individuals currently working (i.e., excluding groups who were students, furloughed, or were not working) who were employed or self-employed. In comparison to individuals employed in ICT occupations, individuals employed in education (27% more likely), health care (29%), social care (43%), transport (19%), retail (23%), hospitality (30%) and manufacturing (18%) were more likely to have tested positive for COVID-19. The analysis also accounts for where individuals are working from. Individuals who are unable to work from home were 30% (OR = 1.30, 95% CIs = 1.23-1.38) more likely to have tested positive for COVID-19 than compared to individuals who were working from home.

**Table 2:** **Model summary for analysing COVID-19 risk by socio-demographic features including occupational group for adults who work. Note: Model also adjusted for time (month). Results placed in bold to emphasise associations where 95% confidence intervals (CIs) do not contain 1**.

| Variable | Odds Ratio | Lower CI | Upper CI |
| --- | --- | --- | --- |
| Male | Reference |  |  |
| Female | 0.958 | 0.909 | 1.009 |
| Age (z-score) | **0.724** | **0.578** | **0.908** |
| Age-squared (z-score) | 1.235 | 0.941 | 1.622 |
| White British | Reference |  |  |
| Any other ethnic group | 1.076 | 0.932 | 1.242 |
| Any other white background | 0.975 | 0.875 | 1.086 |
| Chinese | 0.718 | 0.510 | 1.010 |
| Indian | 1.111 | 0.959 | 1.287 |
| Pakistani | **1.749** | **1.368** | **2.237** |
| Black-African | 1.276 | 0.970 | 1.678 |
| Black Afro-Caribbean | 1.039 | 0.723 | 1.494 |
| Mixed-White & Asian | 0.894 | 0.650 | 1.229 |
| Mixed-White & Black | 1.124 | 0.834 | 1.515 |
| White-Irish | 1.065 | 0.855 | 1.327 |
| ICT | Reference |  |  |
| Teaching and education | **1.274** | **1.135** | **1.429** |
| Health care | **1.285** | **1.140** | **1.450** |
| Social care | **1.428** | **1.207** | **1.689** |
| Transport (incl. storage, logistic) | **1.192** | **1.025** | **1.386** |
| Retail sector (incl. wholesale) | **1.230** | **1.081** | **1.401** |
| Hospitality (e.g. hotel, restaurant) | **1.300** | **1.092** | **1.547** |
| Food production, agriculture, farming | 0.936 | 0.743 | 1.179 |
| Personal services (e.g. hairdressers) | 1.035 | 0.803 | 1.334 |
| Financial services incl. insurance | 1.062 | 0.940 | 1.200 |
| Manufacturing or construction | **1.176** | **1.042** | **1.326** |
| Civil service or Local Government | 1.118 | 0.980 | 1.274 |
| Armed forces | 0.998 | 0.636 | 1.566 |
| Arts, Entertainment or Recreation | 1.022 | 0.851 | 1.227 |
| Other occupation sector | 1.037 | 0.928 | 1.159 |
| Work from home | Reference |  |  |
| Working somewhere else (not your home) | **1.302** | **1.231** | **1.376** |
| Both (from home and somewhere else) | **0.890** | **0.817** | **0.970** |
| Have travelled abroad recently | **1.147** | **1.072** | **1.227** |
| Household size (z-score) | **1.088** | **1.064** | **1.113** |
| Random effects | Variance | SD |  |
| ID (participant) | 2.943 | 1.716 |  |
| Geographical area | 0.147 | 0.384 |  |

The model also finds similar associations for age, household size and whether an individual had travelled abroad as reported previously. Fewer associations were detected by ethnic group, although individuals of Pakistani ethnicity were 75% (OR = 1.75, 95% CIs = 1.37-2.24) more likely to have tested positive than compared to individuals of White British ethnicity.

Plotting the condition mean estimated from the modelled random effect in both models (see Appendix Figure C), allows us to estimate geographical inequalities influencing the likelihood of individuals testing positive for COVID-19. Larger positive values were reported in the North of England especially Liverpool and Manchester, as well as in London. It would suggest that individuals in these regions, often characterised by densely populated areas with higher levels of deprivation, were more likely to have tested positive for COVID-19. Negative values were found in the South West and East of England, regions characterised by rural areas, suggesting that individuals resident in these areas were less likely to have tested positive for COVID-19.

### Estimating change over time in COVID-19 prevalence by work status

The final section of analysis extends the regression analysis presented in the above section to consider how the relationships and associations vary by month. This is achieved through introducing a series of interaction effects into the model. As such, it can be difficult to interpret the results. To aid the interpretation, we calculated the predicted probability (presented as percentages to aid interpretation) of each occupational measure testing positive for COVID-19 for each month by sex (adjusting for other covariates in the model). The models are supplemented through additional analyses stratifying the model by age group (defined as individuals aged less than 40 years, and individuals aged 40 years and over).

First, we consider the work status of all adults (Figure 6). Increasing predicted probability of COVID-19 is observed through the period for all groups other than students, who experienced a higher predicted probability in October before declining thereafter and remaining flat over the remaining months of the study period. The highest predicted probability for each month was otherwise predicted for furloughed individuals, with individuals who were not working having the lowest probability in each month. Stratifying the analyses by age group suggests few differences in estimated risk across each work status group due to the wide uncertainty in estimates with overlapping confidence intervals (Appendix Figure D). The analysis does reveal the higher than expected prevalence of COVID-19 predicted for both furloughed males under 40 years and females under 40 years not working in January.

**Figure 6:**
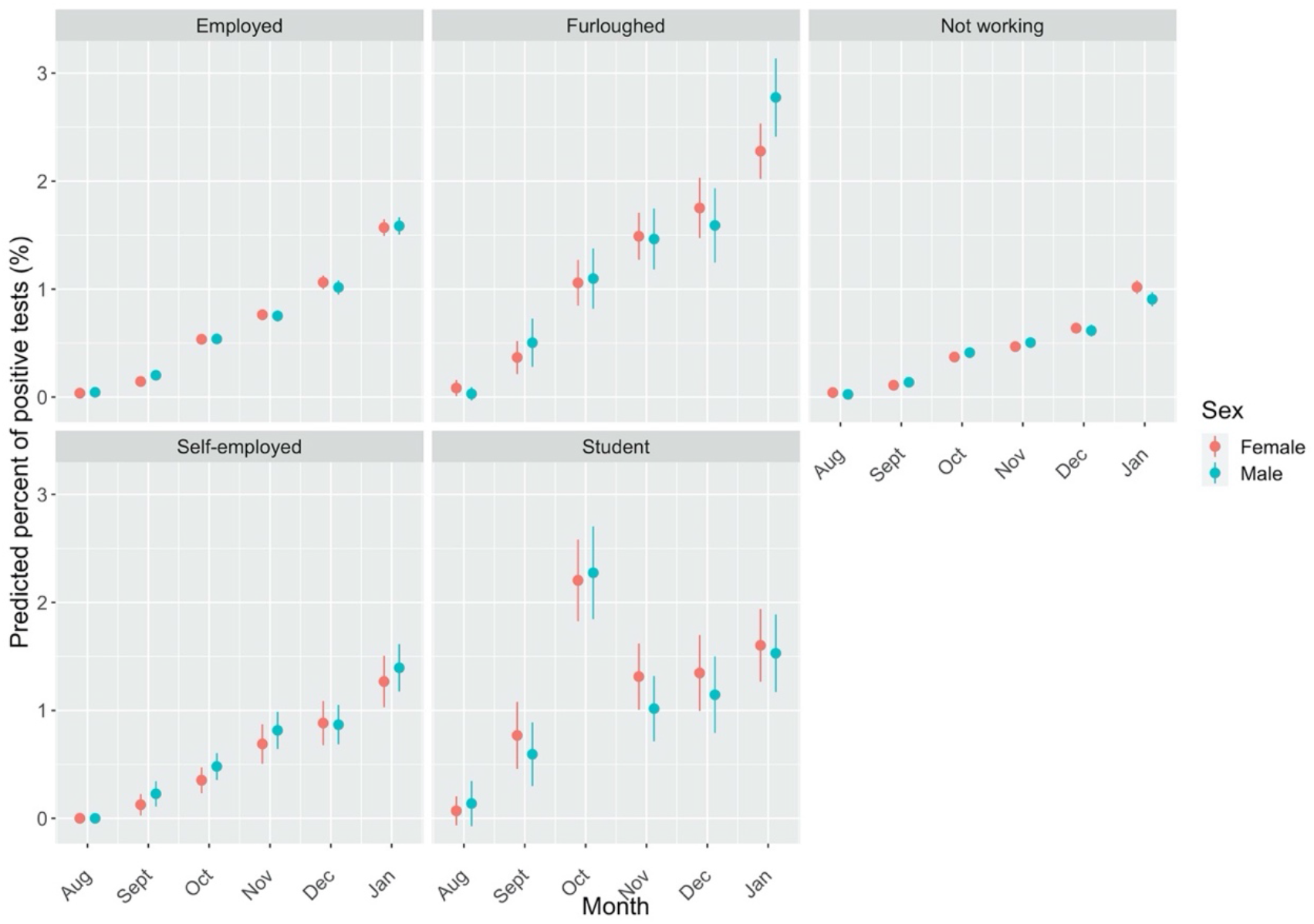
**Predicted probability of testing positive for COVID-19 by work status, sex and month. Note: Estimates are adjusted for geographical location, age, ethnicity, household size and whether an individual had travelled abroad**.

Figure 7 presents the next model, analysing the predicted probability of COVID-19 by work sector among adults who were currently working. A similar trend of higher predicted probability is observed over time and is consistent by work sector. Differences in the predicted probability of COVID-19 between work sector largely follow the results presented in Table 2, with the highest predicted probabilities in January observed for individuals working in transport, hospitality, retail health and social care. One noticeable difference to the general trend is the higher probability in September for women employed in personal services. Stratifying by age group (Figure E) suggests this additional risk is concentrated among women less than 40 years (predicted value = 1.23%, 95% CIs = 0.14% - 2.32%). Similarly, this group observes a large jump in the general trend in December (predicted value = 1.74%, 95% CIs = 0.61% - 2.83%) compared to other sectors (additionally concentrated among younger adults).

**Figure 7:**
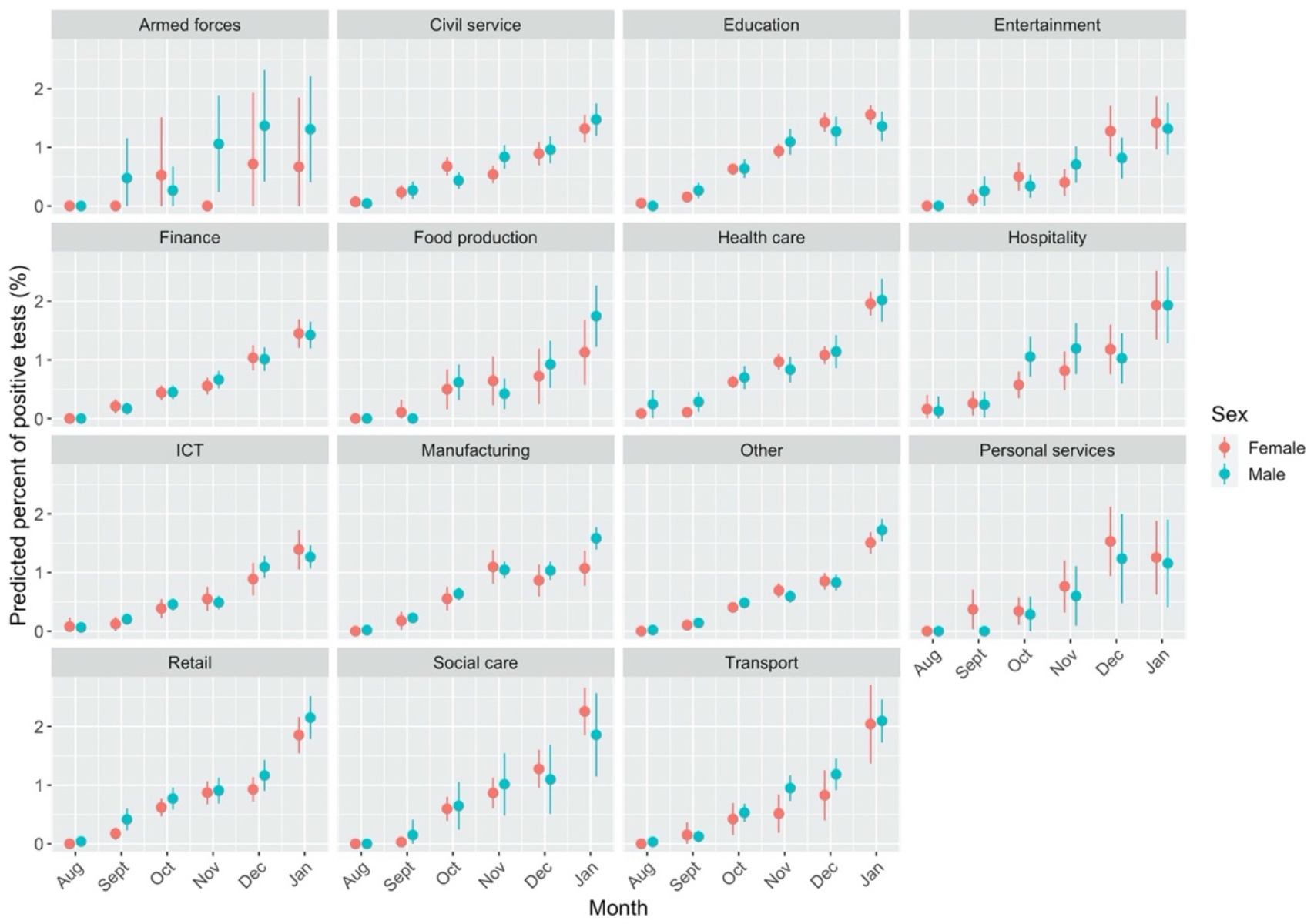
**Predicted probability of testing positive for COVID-19 by work status, sex and month. Note: Estimates for geographical location, age, ethnicity, household size, work location and whether an individual had travelled abroad**.

## Discussion

Our study presents one of the most detailed investigations into the extent of occupational inequalities in COVID-19 for England. For all adults, individuals who were furloughed (i.e., temporarily not working) or students had higher likelihood of COVID-19. Focusing on adults currently working, individuals employed in health or social care, retail, personal services, transport, hospitality and teaching had higher likelihood of testing positive for COVID-19. Likelihood of COVID-19 also varied geographically, with higher risk in densely populated areas across Northern England and London. We also find demographic inequalities with COVID-19 prevalence being higher among younger populations and Pakistani, Black African and Indian ethnic groups. Our findings demonstrate the need for adequate strategies to tackle the social determinants of COVID-19 to equitably manage the pandemic.

The finding that COVID-19 was high among furloughed populations may initially feel counter-intuitive, since individuals may have found it easier to socially distance or isolate compared to those groups employed. While furloughed populations may have fewer work social contacts, evidence suggests that their leisure and social contacts remain higher than other groups (Bridgen, Jewell and Read, no date). It suggests a ‘double jeopardy’ effect whereby individuals are not just negatively impacted by being furloughed (e.g., economic hardship from lost labour opportunities or stress from fear of eventual unemployment (Witteveen, 2020)), but are also more likely to develop COVID-19 that may doubly disadvantaging their health and wellbeing. With the prospect of many furloughed individuals being made permanently unemployed following the end of the furlough system, greater support for these populations will be key to minimise future health inequalities resulting from loss of income (Whitehead, Taylor-Robinson and Barr, 2021).

A similar explanation for the importance of social mixing can help to explain the high prevalence among students. The large spike in prevalence observed in October coincides with the start of most University terms where social mixing of individuals from different regions would have occurred. University student migration represents the largest annual internal migration flow in England (Duke-Williams, 2009), and managing the process safely will be important to minimising further outbreaks.

Among working individuals, we find that COVID-19 prevalence was not equitably spread across work sectors. Higher prevalence of COVID-19 was not just witnessed in patient or care focused professions (e.g., health or social care sectors). We also found that COVID-19 was more common among individuals in work sectors characterised by roles less able to work from home or with greater exposure due to social mixing (e.g., transport, hospitality, retail, personal services or teaching). Our findings follow similar evidence for older adults (50-64 years) on the higher risk of COVID-19 and severe outcomes for key workers (Mutambudzi *et al*., 2021), patterns in national testing records (de Gier *et al*., 2020), as well as for occupational inequalities in COVID-19 mortality (Chen *et al*., 2021; ONS, 2021a). Importantly, we add to this literature through tentatively demonstrating how occupational inequalities were not consistent over time, with tentative evidence of ‘seeding’ of COVID-19 transmission among individuals (particularly young females) employed in personal services (e.g., hairdressers) at the start of the second wave.

Our findings suggest the need for better workplace interventions across diverse roles that can help contain COVID-19 transmission, whilst allowing individuals and employers to continue their social and economic activities (Mutambudzi *et al*., 2021). Occupational roles will need to further adapt to protect their employees from COVID-19. Minimising social contacts or mixing within occupational roles through sufficient preventative measures may be valuable. One study suggested that limiting the number of social contacts at work was the most important strategy for lowering the ‘R’ number if society keeps schools open (Brooks-Pollock *et al*., 2021). Repeat testing of employees may help to manage outbreaks, however testing behaviours can widen inequalities (Green *et al*., 2021). Support for lost earnings if individuals have to self-isolate will be key, especially as some of the work sectors identified here with higher prevalence (e.g., retail or hospitality) are characterised by low wages (Paremoer *et al*., 2021). However, our findings of high prevalence of COVID-19 for furloughed and student populations demonstrates the need for broader strategies than just occupation-related interventions to help manage COVID-19 and tackle the drivers of health inequalities.

Our analyses also demonstrate wide ethnic inequalities in COVID-19 prevalence. Likelihood of having had a positive COVID-19 test was higher among Pakistani, Black African, and Indian groups than compared the majority White British population. Our results follow evidence from other studies and other outcomes relating to COVID-19 (Public Health England, 2020; Harrison *et al*., 2021; HM Government, 2021b). While work sector attenuated these associations for adults currently working and may partly explain higher risk among some ethnic groups, explaining away the differences between ethnic groups through work sectors should not detract from the extent of ethnic inequalities of COVID-19. Ethnicity intersects with occupation, with the social sorting of disadvantaged and marginalised ethnic groups into employment roles that have greater exposure to COVID-19 risk through higher social contacts or inability to work from home (Platt and Warwick, 2020; Paremoer *et al*., 2021). Future research should explore the intersecting pathways between occupation and ethnicity to improve our understanding of why these inequalities exist.

There are several limitations to our study. We do not account for all possible explanatory factors (e.g., deprivation, social distancing behaviours) that may explain occupational inequalities due to a lack of suitable data available for our analysis. Through focusing on work sector, rather than specific occupation or role, we may be limited how generalisable our findings are. For example, teaching and education would include both primary and secondary teachers who were expected to teach classes face-to-face and therefore have different exposures to University lecturers who could far easier adapt to work from home. The lack of specific occupation categories may therefore under-estimate the specific risks and inequalities faced across England. Finally, our analyses are association-based and do not explore potential causal pathways or mechanisms through how and why occupation influences COVID-19 risk. Future research should extend our analyses to consider the specific mechanisms that may explain, mediate or moderate risk.

## Conclusion

Our study, using novel large scale longitudinal data, demonstrates the importance of the social determinants of health through work and occupation in understanding the unequal burden in COVID-19 prevalence. We find complex and diverse pathways through which SARS-CoV-2 transmission may occur across numerous work sectors which can exacerbate, reinforce and create new health inequalities. Population groups employed in sectors with greater social contacts, less able to work from home or having front facing roles have greater likelihood of COVID-19. Additionally, groups that have experienced social and economic harms through furlough appear to have experienced a double jeopardy in greater likelihood of COVID-19. Although our results focuses on the period of the COVID-19 pandemic, they also reflect longer-term societal patterns confirming that COVID-19 has reinforced and amplified existing health inequalities (Marmot *et al*., 2020; Whitehead, Taylor-Robinson and Barr, 2021).

## Data Availability

Data cannot be openly shared, however the raw data are freely available for researchers to apply for access via the ONS Secure Research Service. Details of can be found here https://www.ons.gov.uk/aboutus/whatwedo/statistics/requestingstatistics/approvedresearcherscheme

## Acknowledgements

This work was supported by the Economic and Social Research Council [grant number ES/L011840/1]. This work was produced using statistical data from ONS. The use of the ONS statistical data in this work does not imply the endorsement of the ONS in relation to the interpretation or analysis of the statistical data. This work uses research datasets which may not exactly reproduce National Statistics aggregates.

