## Appendix for "Occupational inequalities in the prevalence of COVID-19: A longitudinal observational study of England, August 2020 to January 2021"

**Table A: Descriptions of work sector categories.**

| Work Sector | Example occupations |
|---|---|
| ICT | IT technician, software engineer, programmer, web designer |
| Education | Teacher, lecturer, school workers |
| Health care | Doctor, nurse, dentists, health professionals, pharmacists |
| Social care | Carer, social workers, welfare professionals |
| Transport | Bus driver, logistics, storage firms, taxi driver |
| Retail sector | Shop assistant, retail cashier, check-out operators |
| Hospitality | Solider, naval officer, air forces pilot |
| Food production | Farmer, agricultural labourer, butcher, baker |
| Personal services | Hairdresser, barber, cleaner, beautician |
| Finance | Insurance, banking, accountant, |
| Manufacturing | Construction, skilled and non-skilled trades, engineers |
| Civil service | Local Government worker, health and safety officer, |
| Armed forces | Solider, naval officer, air forces pilot |
| Entertainment | Actor, artist, musician, recreational officer |

**Table B: Sample summary statistics.**

| Variable | Mean (SD) |
|---|---|
| Age | 53.96 (17.1) |
| **Variable** | **Percentage (%)** |
| *Test outcome* | |
| Negative | 96.9 |
| Positive | 0.9 |
| Void | 2.2 |
| *Sex* | |
| Male | 46.6 |
| Female | 53.4 |
| *Ethnicity* | |
| White British | 87.9 |
| Any other ethnic group | 2.0 |
| Any other white background | 4.1 |
| Chinese | 0.5 |
| Indian | 1.9 |
| Pakistani | 0.4 |
| Black-African | 0.4 |
| Black Afro-Caribbean | 0.3 |
| Mixed-White & Asian | 0.4 |
| Mixed-White & Black | 0.4 |
| White-Irish | 1.7 |

| | |
|---|---|
| *Work status* | |
| Employed | 47.1 |
| Self-employed | 4.3 |
| Furloughed | 3.9 |
| Not working | 42.6 |
| Student | 2.1 |
| *Work sector* | |
| ICT | 7.6 |
| Education | 14.4 |
| Health care | 11.4 |
| Social care | 3.0 |
| Transport | 4.1 |
| Retail sector | 7.5 |
| Hospitality | 3.3 |
| Food production | 1.7 |
| Personal services | 1.4 |
| Finance | 8.7 |
| Manufacturing | 10.0 |
| Civil service | 7.1 |
| Armed forces | 0.4 |
| Entertainment | 2.9 |
| Other | 16.5 |
| *Month* | |
| August | 6.0 |
| September | 12.9 |
| October | 23.1 |
| November | 20.5 |
| December | 18.3 |
| January | 19.2 |

**Table C: Percentage of records with missing data.**

| Variable | Missing (%) |
|---|---|
| Test outcome | 3.57 |
| Age | 0.00 |
| Sex | 0.00 |
| Ethnicity | 0.02 |
| Work status | 0.06 |
| Work sector | 3.57 |
| Month | 0.00 |
| Geography | 10.98 |
| Travel abroad | 3.20 |
| Household size | 0.00 |

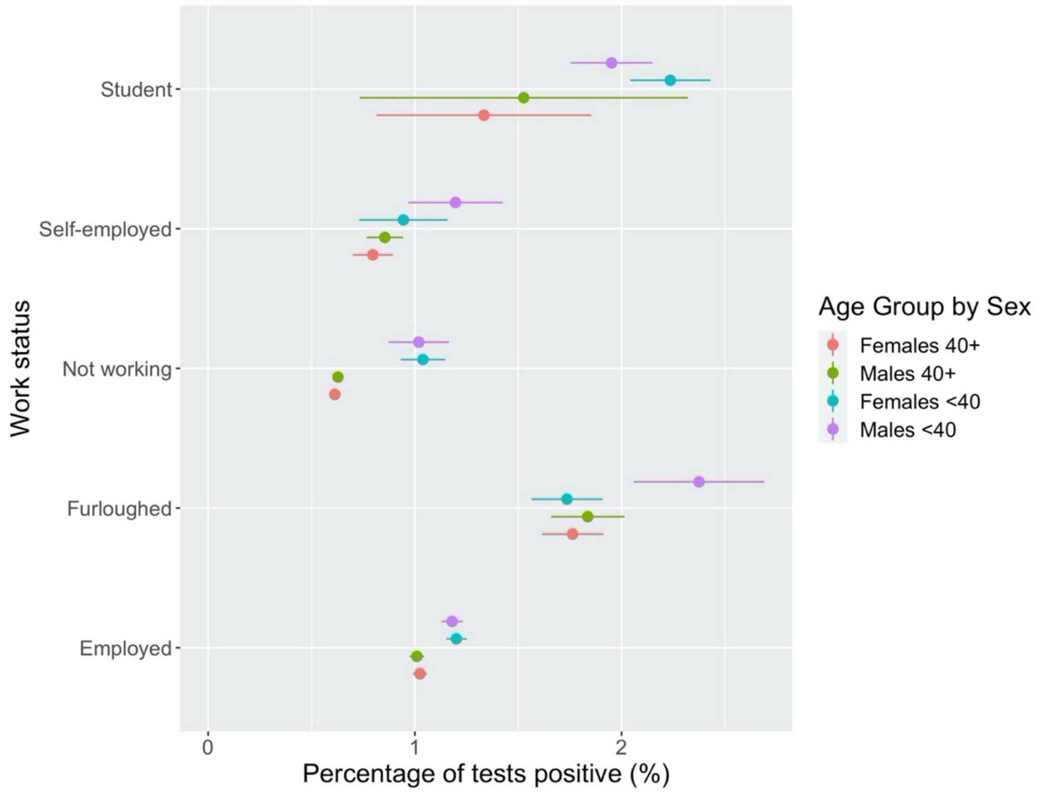


**Figure A: COVID-19 Prevalence by work status and sex for individuals by age group.**

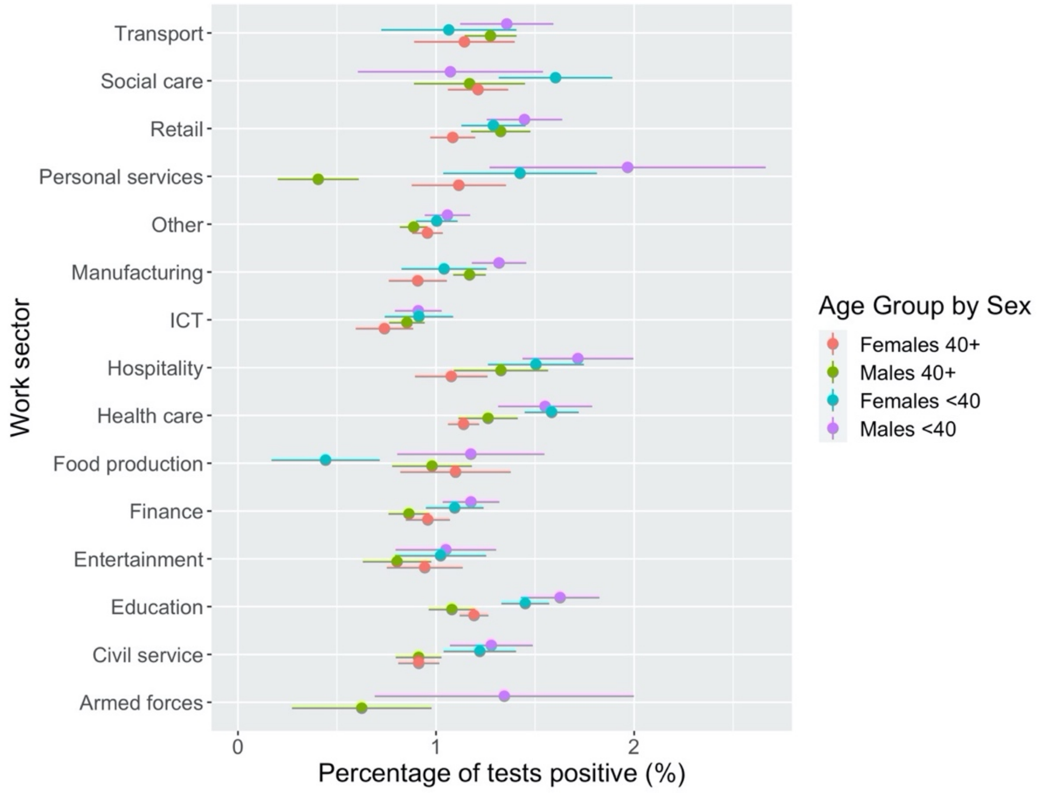


**Figure B: COVID-19 prevalence by work sector and sex for individuals by age group. Note: estimate for females employed in the armed forces excluded due to counts <10 to preserve ONS data disclosure standards.**

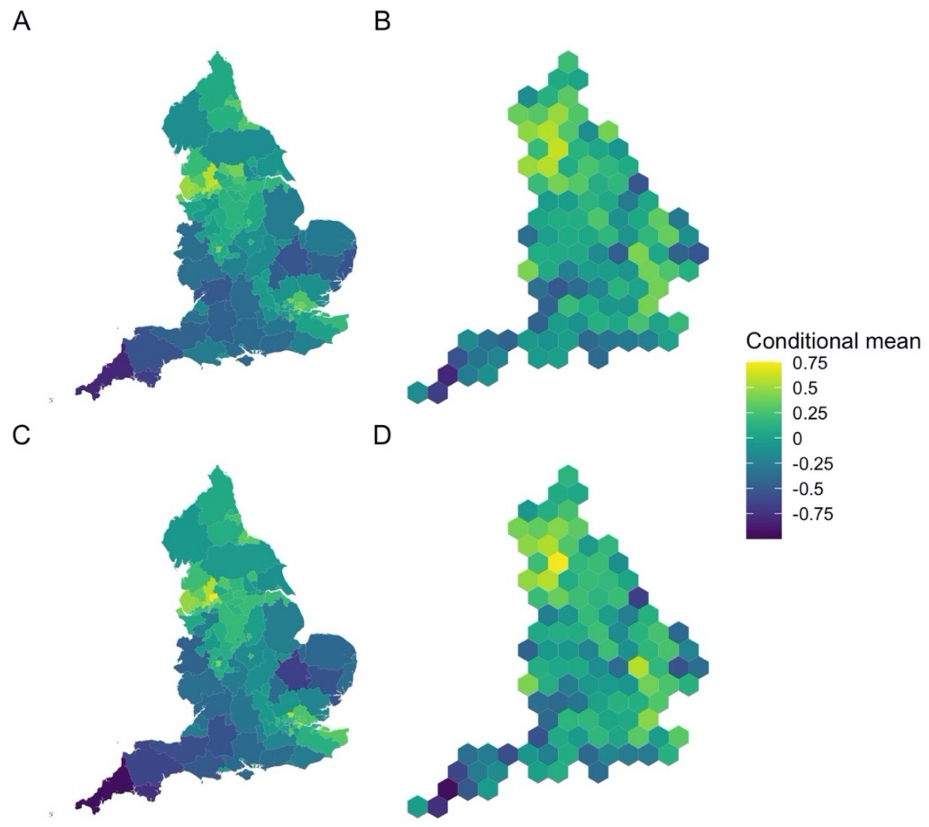


**Figure C: Conditional mean estimated from the overall analytical models. Plot A: Actual geography - Table 1. Plot B: Hex map - Table 1. Plot C: Actual geography - Table 2. Plot D: Hex map - Table 2.**

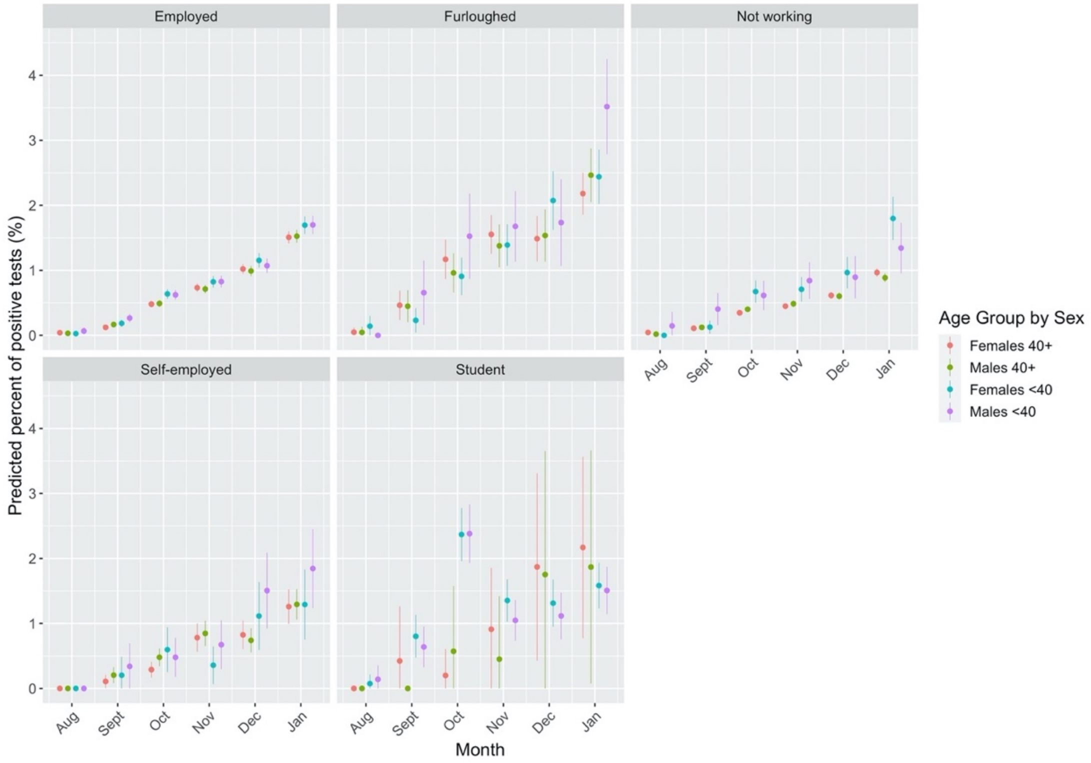


**Figure D: Predicted probability of testing positive for COVID-19 by work status, sex and month stratified by age group.**

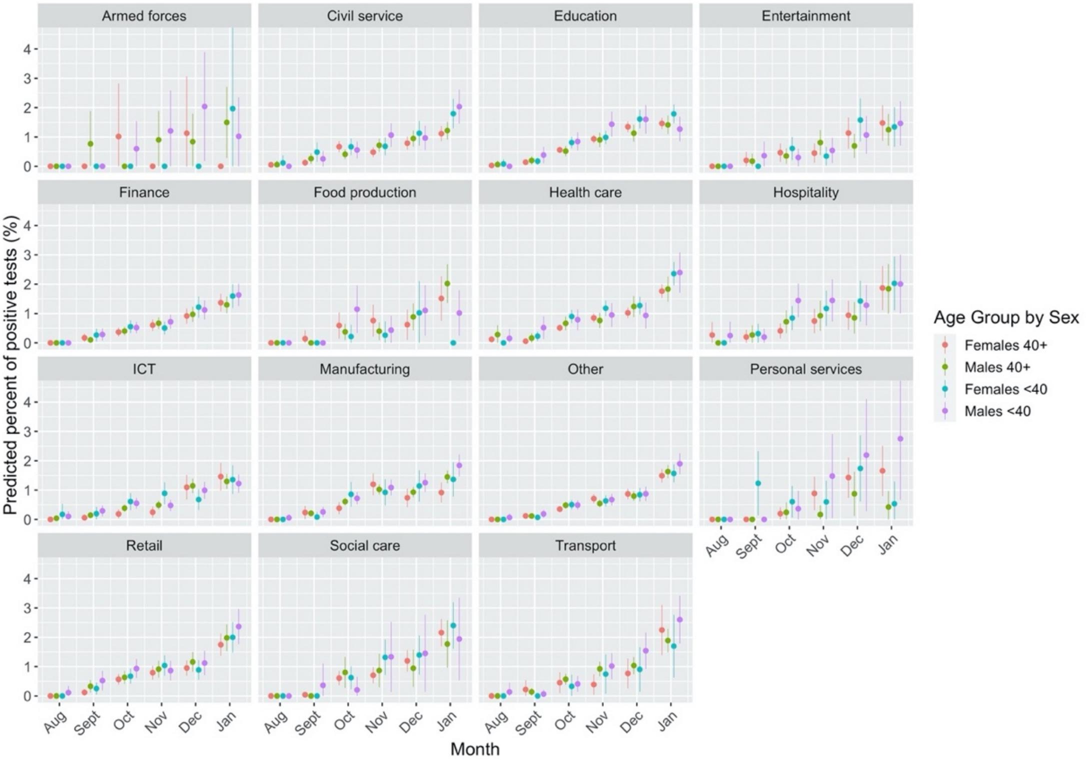


**Figure E: Predicted pry of testing positive for COVID-19 by work sector, sex and month stratified by age group.**
